# Another doubling of excess mortality in the United States relative to its European peers between 2017 and 2021

**DOI:** 10.1101/2022.03.21.22272722

**Authors:** Patrick Heuveline

**Author notes:** **(Corresponding) Author: Patrick Heuveline, PhD**.

## Abstract

A mortality gap between the United States and other high-income nations emerged before the pandemic. International comparisons of Covid-19 mortality suggest this gap might have increased during the pandemic.

Applying average mortality rates of the five largest West European countries to the US population shows that the number of “excess deaths” attributable to this mortality gap continues to increase year after year in the United States. The annual number of such excess deaths has doubled between 2017 and 2021, with most of the increase occurring during the pandemic (+89.1% between 2019 and 2021). In 2021, excess mortality in the United States relative to its European peers contributed 892,491 excess deaths, amounting to 25.8% of all US deaths that year, up from 15.7% in 2017.

Of the 450,224 excess deaths added between 2017 and 2021, 42,317 are attributable to population change (9.4%), 230,672 to differential rates of Covid-19 mortality (51.2%), and the remaining 177,235 to differential rates of mortality from other causes (39.4%, possibly including misclassified deaths due to Covid-19). The contribution of Covid-19 mortality to excess mortality in the United States (relative to its European peers) grew between 2020 and 2021 due to diverging trends in Covid-19 mortality, especially towards the end of 2021 as US vaccination rates plateaued at lower levels than in European countries. While this contribution might be transient, divergent trends in mortality from other causes persistently separates the United States from West European countries. Excess mortality is particularly high between ages 15 and 64. In 2021, nearly half of all US deaths in this age range are excess deaths (48.0%).

---

The pre-pandemic emergence of a gap in mortality between the United States and other high-income nations is well documented. Arguably the most salient measure of that mortality gap is the number of “excess deaths.”. This number is estimated by subtracting from the actual number of deaths a counterfactual number of deaths obtained after replacing the prevailing sex- and age-specific mortality rates with more favorable rates. In recent months, the concept has been primarily used to assess the impact of the pandemic by substituting (lower) pre-pandemic rates for the actual rates. But the concept has long been used to assess the consequences of mortality differentials whether between population sub-groups, e.g., racial/ethnic differentials, ^1^ or between populations. ^2^

A previous study tracked the annual number of excess deaths in the United States from 2000 to 2017 by estimating how many fewer deaths would have occurred had the country faced the same mortality rates as a composite of the five largest Western European countries (England and Wales, France, Germany, Italy, and Spain). With a combined population size very similar to that of the United States, these five countries provide a realistic benchmark for a large, diverse, and wealthy nation. The results were striking: the annual number of excess deaths estimated so nearly doubled from 226,165 in 2000 to 400,732 in 2017, amounting to one in seven deaths that year (14.2%). ^3^ But this gap can be expected to have continued to expand during the pandemic, as provisional data suggested the United States experienced larger mortality declines than peer countries. ^4^ Indeed, the difference in Covid-19 mortality between the United States and the same five countries has been estimated to add 132,173 excess deaths in 2020, growing to 190,867 excess deaths for the 12-month period from April 1, 2020 to April 1, 2021. ^5^

Using both all-cause and Covid-19 mortality data up to end of 2021, this study estimates the annual number of US excess deaths relative to the same five European countries from 2017 to 2021, and the specific contribution of differences in Covid-19 mortality from 2019 to 2021.

## Material and methods

Excess deaths are estimated by comparing actual counts of deaths by sex and five large age groups (0- 14, 15-64, 65-74, 75-84 and 85+) in the United States with counterfactual numbers of deaths. The counterfactual numbers are derived by applying population-weighted average sex- and age-specific European rates to the US size of these sex- and age-groups each year. To assess the contribution of population changes, another set of counterfactual numbers is derived by holding the US sex and age groups to their 2017 size. To assess the specific contribution of Covid-19 mortality, counterfactual numbers of Covid-19 deaths in the United States are estimated for each calendar year, 2020 and 2021, and for three twelve-month periods in-between. All data are from publicly available sources, see Supplementary Information for more methodological details.

## Results

Based on the weighted average of the rates for the five European countries, excess deaths are estimated to have doubled to 892,491 in 2021 (25.8% of all deaths that year) from 442,267 in 2017 (15.7% of all deaths that year). (2017 figures differ from those estimated in an earlier study^3^ due to the revision of intercensal population estimates following the 2020 Census, see Supplementary Information for details).

Holding US age groups at their 2017 sizes shows that population changes only accounted for a small fraction of the 2017-2021 increase in excess deaths (42,317 excess deaths, 9.4% of the 2017-2021 increase).

More than half of the 2017-2021 increase in excess deaths (51.2%) can be attributed to international differences in Covid-19 mortality. The number of US deaths attributed to Covid-19 that can be considered as excess deaths (relative to its European peer countries) increased from 121,366 in 2020 to 230,672 in 2021 (Table 1). This dynamic is further described in Figure 1 which shows the total number of deaths attributed to Covid-19 and excess deaths among those for 2020, 2021, and three intermediate 12-month periods in-between. The number of US deaths attributed to Covid-19 that are excess deaths increased despite a modest decline in the total number of US deaths attributed to Covid-19 in a 12- month period (69,582 fewer deaths in 2021 than in the first 12 months of pandemic mortality, from April 1^st^, 2020 to April 1^st^, 2021). This modest decline was more than compensated by an increase in the fraction of excess deaths among those attributed to Covid-19. Relatively stable until mid-2021 (from 34.6% for 2020 to 37.2% for the mid-2020 to mid-2021 period), that fraction decreased to 29.2% in the 12-month period ending on October 1st, 2021, before bouncing back to 48.2% for the calendar year 2021.

**Table 1:** Annual estimates of excess mortality in the United States (number of excess deaths and excess-to-all-death ratios) and contributions to 2017-2021 change

|  | Year |  |  |  |  |
| --- | --- | --- | --- | --- | --- |
|  | 2017 | 2018 | 2019 | 2020 | 2021 |
| Total number of excess deaths | 442,267 | 433,811 | 471,992 | 745,109 | 892,491 |
| Excess-to-all-death ratios | 15.7% | 15.3% | 16.5% | 22.2% | 25.8% |
| Total number with 2017 population | 442,267 | 427,968 | 459,122 | 716,455 | 850,174 |
| Ratios with 2017 population | 15.7% | 15.4% | 16.7% | 22.3% | 25.8% |
| Contribution of population change |  | 5,843 | 12,870 | 28,654 | 42,317 |
| Share of change in excess deaths |  | -69.1% | 43.3% | 9.5% | 9.4% |
| Contribution of Covid-19 mortality |  |  |  | 121,366 | 230,672 |
| Share of change in excess deaths |  |  |  | 40.1% | 51.2% |
| Contribution of other-cause mortality |  | -14,299 | 16,855 | 152,822 | 177,235 |
| Share of change in excess deaths |  | 169.1% | 56.7% | 50.5% | 39.4% |

Notes: Author's calculations from the Human Mortality Database, Short-Term Fluctuation Series (STFS), with population-weighted average of European countries as counterfactual. See Supplementary File for estimation details.

**Figure 1.**
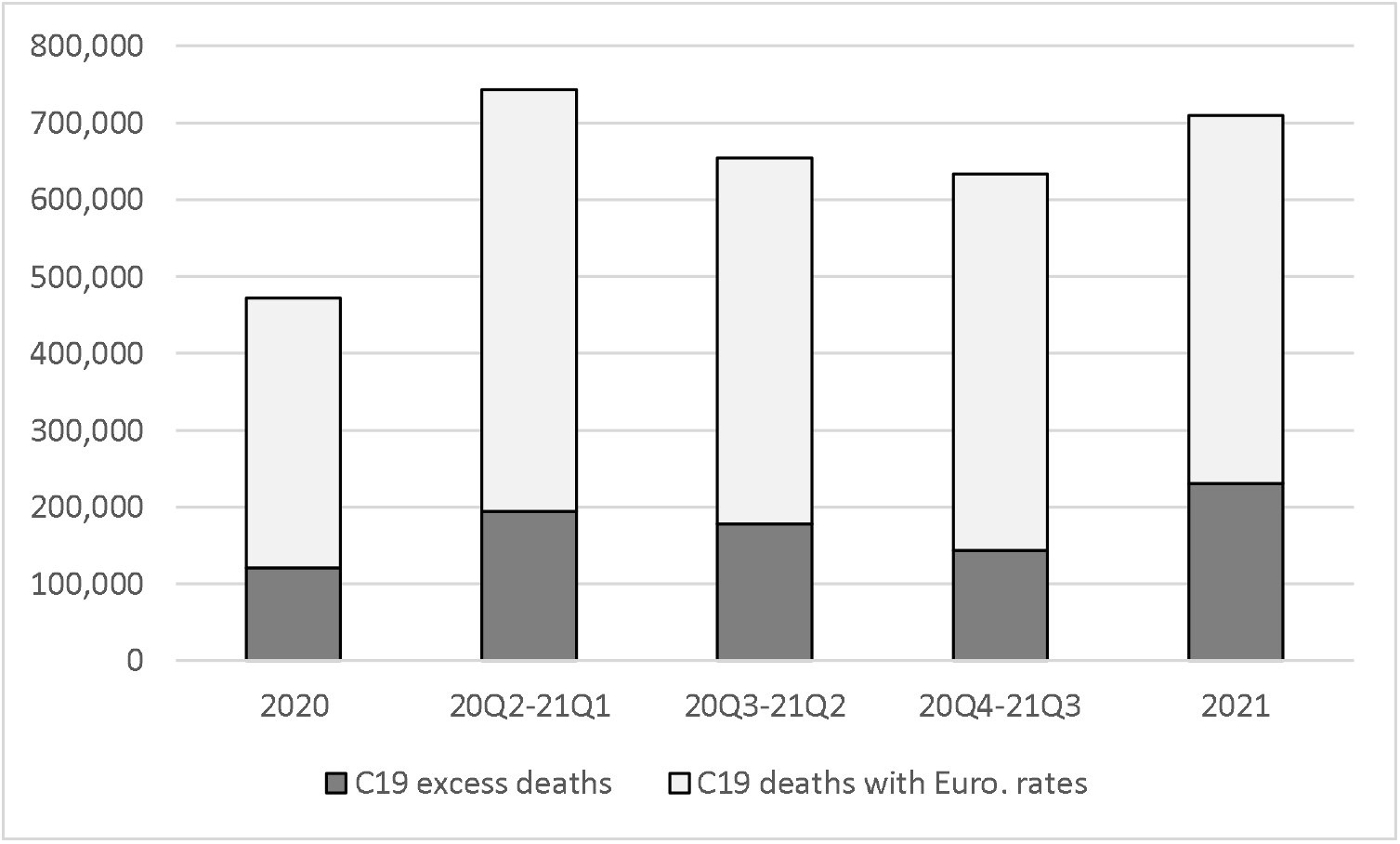
Number of US deaths attributed to Covid-19 by 12-month period, broken down as the sum of Covid-19 deaths estimated by applying the population-weighted average of the European death rates from Covid-19 to the US population (light grey) and the excess deaths from Covid-19 (dark grey). The first (left) and last (right) periods are calendar years. The three intermediate 12- month period are shifted by one quarter each: i.e., 20Q2-21Q1 refers to the period from April 1^st^, 2020 to April 1^st^, 2021.

Table 2 shows the distribution of 2021 excess deaths by sex and large age groups as well as the proportion of deaths in each sex and age-group that can be considered excess deaths. In 2021, 31.0% of all excess deaths (both sexes combined) occurred to males between the ages of 15 and 64 and another 19.1% to females in the same age range. Altogether, more than half of the excess deaths were in that age range in 2021 (50.1%). Conversely, nearly half of all deaths between the ages of 15 and 64 are excess deaths, a higher proportion than in other age groups, and the proportion is even higher for females (49.6%) than for males (46.4%).

**Table 2:** Excess US deaths, and ratios to all US deaths, 2021, by sex and age group

|  | 0-14 | 15-64 | 65-74 | 75-84 | 85+ | All ages |
| --- | --- | --- | --- | --- | --- | --- |
| Females | 5,685 | 170,268 | 112,059 | 94,141 | 53,944 | 436,097 |
| Males | 7,253 | 276,648 | 97,449 | 53,588 | 21,457 | 456,394 |
| Both sexes | 12,938 | 446,916 | 209,508 | 147,729 | 75,401 | 892,491 |
| Distribution of all-age, both-sex excess deaths |  |  |  |  |  |  |
| Females | 0.6% | 19.1% | 12.6% | 10.5% | 6.0% | 48.9% |
| Males | 0.8% | 31.0% | 10.9% | 6.0% | 2.4% | 51.1% |
| Both sexes | 1.4% | 50.1% | 23.5% | 16.6% | 8.4% | 100.0% |
| Excess-to-all-death ratio in sex and age-group |  |  |  |  |  |  |
| Females | 43.1% | 49.6% | 36.8% | 23.8% | 9.6% | 26.9% |
| Males | 44.0% | 46.4% | 23.3% | 12.4% | 5.8% | 24.9% |
| Both sexes | 43.6% | 48.0% | 29.0% | 17.8% | 8.1% | 25.8% |

## Discussion

These results show that the impressive increase in the number of excess deaths in the United States relative to its peer West European countries between 2000 and 2017 (77.2%) ^3^ was followed by an even larger increase, a doubling in only four years (2017-2021). While death data by single year of age are not yet available to fully replicate the analysis from 2017 up to the present, data by large age groups provide a close approximation and demonstrate an unambiguous trend. From 2017 to 2019, excess deaths increased by another 6.7%, bringing the cumulative increase between 2000 and 2019 to 89.1%. From 2019 to 2021, excess deaths increased by the same proportion (89.1%) as in the previous 19 years, to approach 900,000 in 2021 (892,491 deaths). The proportion of excess deaths among all deaths also surged from more than one in seven in 2017 (15.7%) to more than one in four in 2021 (25.8%).

As between 2000 and 2017, population change (growth and aging) played a relatively small role in the increase since 2017. International differences in rates of death attributed to Covid-19 explains half of the 2017-2021 increase in excess deaths. These differences might be linked in part to differences in the prevalence of comorbidities associated with Covid-19 case-fatality rate. Through mid-2021, differences in Covid-19 death rates remained relatively stable.^5^ For the second half of the year, the gap in Covid-19 death rates first narrowed as vaccines became available to the general population earlier in the United States than in Europe except for England and Wales. By the end of 2021, vaccination rates in the United States were plateauing at lower levels than those achieved in the West European countries, ^6^ however, and the contribution of Covid-19 mortality to overall excess mortality grew larger in 2021 than it had been in previous 12-month periods since the beginning of the pandemic.

Differences in mortality from other causes as well also increased, contributing an additional 177,235 excess deaths between 2017 and 2021 (39.4% of the total 2017-2021 increase). Mortality from some causes might be expected to improve (e.g., from reduction in motor-vehicle traffic) ^7^ as well as to worsen for other causes (e.g., because of the impact of the pandemic on the quality of hospital care) ^8^ during the pandemic. While the net effect has been a substantial reduction in deaths from other causes in the five European countries, partially compensating the additional deaths directly attributable to Covid-19, ^9^ the United States has experienced an overall increase in deaths from other causes. ^10^ These might reflect in part a relatively higher degree of Covid-19 deaths being attributed to other causes, ^11^ but rates of death from unintentional injuries have also been increasing in the United States, ^12^ in particular those involving synthetic opioids^13^ or alcohol^14^. While the contribution of Covid-19 should tilt excess mortality towards older ages, adult deaths between the ages of 15 and 64 thus continue provide the majority excess deaths (50.1%) and the share of deaths that are excess deaths is largest between these ages (49.6% for females, 46.4% for males).

The dramatic surge in excess mortality in the United States during the pandemic is, in Eileen Crimmins’ words, “an acute demonstration of our chronic problem.” ^15^ So acute that the proportion of US adult deaths that were “excess deaths” in 2021 (48% for both sexes combined between ages 15 and 64) far exceeds the fraction of US deaths at ages 50 and over that were attributable to smoking in 2003 (24%), ^16^ or the fraction of adult deaths (ages 20-64) attributable to alcohol in Russia in 2002 (29%). ^17^

## Data Availability

All materials from other sources are publicly available.

## Statements

### Funding Statement

The author benefited from facilities and resources provided by the California Center for Population Research at UCLA (CCPR), which receives core support (P2C-HD041022) from the Eunice Kennedy Shriver National Institute of Child Health and Human Development (NICHD).

### Conflict of Interest Disclosure

The author has no conflict of interest.

### Ethics Approval Statement

The study has no human subjects. All data are publicly available and contain no identifying information.

### Patient Consent Statement

The study was conducted without patient involvement. Patients were not invited to comment on the study design and were not consulted to develop patient relevant outcomes or interpret the results. Patients were not invited to contribute to the writing or editing of this document for readability or accuracy.

### Data Statement

All materials from other sources are publicly available.

## Notes

### Competing Interest Statement

The authors have declared no competing interest.

### Summary of Updates

Writing update (analyses unchanged)

## References

1. Wrigley-Field, E. (2020). US racial inequality might be as deadly as COVID-19. Proc Natl Acad Sci U S A., 117(36) 21854–21856. https://doi.org/10.1073/pnas.2014750117

2. Heuveline, P., Guillot, M. & Gwatkin, D. (2002). The uneven tides of the epidemiological transition. Soc Sci Med 55(2) 313–322. https://doi.org/10.1016/s0277-9536(01)00172-1

3. Preston, S. H. & Vierboom, Y. C. (2021). Excess mortality in the United States in the 21^st^ century. Proc Natl Acad Sci U S A., 118, Article e2024850118. https://www.pnas.org/doi/10.1073/pnas.2024850118

4. Woolf, S. H., Masters, R. K., & Aron, L. Y. (2021). Effect of the covid-19 pandemic in 2020 on life expectancy across populations in the USA and other high income countries: Simulations of provisional mortality data. BMJ 373:n1343. https://doi.org/10.1136/bmj.n1343

5. Heuveline, P. (2022). The COVID-19 pandemic adds another 200,000 deaths (50%) to the annual toll of excess mortality in the United States. Proc Natl Acad Sci U S A., 118, Article e2107590118. https://www.pnas.org/doi/10.1073/pnas.2107590118

6. Our World In Data (OWID). Coronavirus (COVID-19) vaccinations. https://ourworldindata.org/covid-vaccinations

7. Yasin, Y. J., Grivna, M. & Abu-Zidan, F. M. (2021). Global impact of covid-19 pandemic on road traffic collisions. World J Emerg Surg, 16, Article 51. https://doi.org/10.1186/s13017-021-00395-8

8. Mansfield, K. E., Mathur, R., Tazare, J. et al. (2021). Indirect acute effects of the COVID-19 pandemic on physical and mental health in the UK: a population-based study. Lancet Digit Health, 3, E217-E230. https://doi.org/10.1016/S2589-7500(21)00017-0

9. Pison, G. & Meslé, F. (2021). France 2020: 68,000 excess deaths attributable to COVID-19. Population & Societies, 587. https://www.ined.fr/en/publications/editions/population-and-societies/france-2020-68000-additional-deaths-attributable-covid-19-epidemic/

10. Ahmad, F. B. & Anderson, R. N. (2021). The leading causes of death in the US for 2020. JAMA, 325, 1829–1830. https://doi.org/10.1001/jama.2021.5469

11. Ackley, C. A., Lundberg, D. J., Ma, L., Elo, I. T., Preston, S. H. & Stokes, A. C. (2022). County-level estimates of excess mortality associated with COVID-19 in the United States. SSM Popul Health, 17, Article 101021. https://doi.org/10.1016/j.ssmph.2021.101021

12. Arias, E., Tejada-Vera, B., Kochanek, K. D., & Ahmad, F. B. (2022). Provisional life expectancy estimates for 2021. Vital Stat Rapid Release 23. Hyattsville, MD: National Center for Health Statistics. https://dx.doi.org/10.15620/cdc:118999

13. National Institute on Drug Abuse. Overdose death rates. Accessed April 19, 2022. https://nida.nih.gov/drug-topics/trends-statistics/overdose-death-rates

14. White, A. M., Castle, I.-J. P., Powell, P. A., Hingson, R. W. & Koob, G. F. (2022). Alcohol-related deaths during the COVID-19 pandemic. JAMA 327(17):1704–1706. https://doi.org/10.1001/jama.2022.4308

15. Crimmins, E. (2021) Life expectancy and health expectancy in the 21^st^ century: the unthinkable, the inconceivable, and the unknowable. PAA Presidential Address 2021. https://www.youtube.com/watch?v=yltaGA4GpNE&t=1364s.

16. Preston, S. H., Glei, D. A. & Wilmoth, J. R. (2010). A new method for estimating smoking-attributable mortality in high-income countries. Int J Epidemiol, 39, 430–438. https://doi.org/10.1093/ije/dyp360

17. Rehm, J., Sulkowska, U., Mańczuk, M., Boffetta, P., Powles, J., Popova, S., Zatoński, W. (2007). Alcohol accounts for a high proportion of premature mortality in central and eastern Europe. Int J Epidemiol, 36, 458–467. https://doi.org/10.1093/ije/dyl294

