## Supplementary material for "Another doubling of excess mortality in the United States relative to its European peers between 2017 and 2021": Data and methods supplement

**Supplementary Information: Extended Material and Methods**

This study first estimates excess deaths from all causes in the United States from the Human Mortality Database’s Short-Term Mortality Fluctuation (STMF).^1^ These data include death counts and death rates for every week up to the end of 2021 in France, Germany, Italy, Spain, the United Kingdom, and the United States, by sex and five large age groups (0-14, 15-64, 65-74, 75-84 and 85+). To compare with an earlier estimate for 2017 that benefited from disaggregated data by single year of age^2^ (still unavailable for 2020 and 2021 at this writing), excess deaths were first re-estimated for 2017. Two sets of annual estimates were derived. The first one used as counterfactual death rates for the US the arithmetic (unweighted) average of the year, sex, and age-group specific death rates in each of the five European countries. The second one used a population-weighted average, that is, deaths and population size for each year, sex, and age-group in the five European countries were added separately, and a corresponding death rate was obtained by dividing total deaths by total population size.

For 2017, the only year for which the comparison is possible with the data currently available, estimates based on the STMF data retrieved in January 2022 were quite close to the estimate based on single-year-of-age data (400,732 excess deaths): 389,305 excess deaths with the weighted average and 412,232 excess deaths with the arithmetic average of the five European rates. Results from the 2020 Census, however, led to a revision of intercensal population estimates in April 2022. Numbers of older adults were revised downward in particular, leading to an increase in the death rates at older ages. With these updated US death rates, excess-death estimates increased (to 442,267 and 465,069 deaths) and are no longer comparable to the original estimate based on single-year-of-age data.

Excess deaths were then calculated for each year from 2017 to 2021 using the weighted average of the European rates. To assess the contribution of population changes to the 2017-2021 trend in excess deaths, another set of annual estimates was derived by holding the US sex and age groups to their 2017 size. To assess the specific contribution of Covid-19 mortality in 2020 and 2021 and its dynamics, counterfactual numbers of Covid-19 deaths in the United States were estimated for each calendar year, 2020 and 2021, as well as for the three four-quarter periods in between (ending in the first, second and third quarter of 2021 respectively). Because countries report their Covid-19 deaths by age using different age groupings,^3^ the previous approach was modified as follows.

The US sex- and age-specific rates of death attributed to Covid-19 for each period was derived from Covid-19 death counts from the Centers for Disease Control and Prevention’s National Center for Health Statistics (NCHS)^4^ and mid-year US population estimates by age and sex from the United Nations (UN) Population Division.^5^ These rates were then combined with mid-year population estimates by age and sex from the UN to estimate a counterfactual number of Covid-19 deaths for France, Germany, Italy, Spain, the United Kingdom and the United States in each of these periods. Comparative Mortality Ratios (CMR) of deaths attributed to Covid-19, as reported on the Johns Hopkins University (JHU) dashboard,^6^ to the corresponding counterfactual numbers above were then calculated for the US and the aggregate of the five European countries in each period. Due to slight differences between the US numbers reported by NCHS and JHU, the US CMR are not exactly equal to unity. To account for these reporting differences, the European CMRs was divided by the corresponding US CMR in each period. This CMR ratio was taken to represent the fraction of deaths attributed to Covid-19 deaths in the United States that would have occurred in the absence of international differences in Covid-19 mortality and completeness of reporting Covid-19 deaths. The remainder are considered to represent the share of deaths attributed to Covid-19 in each period that are excess deaths (relative to the five European countries).

**References**

1. Jdanov, D.A., Alustiza-Galarza, A., Shkolnikov, V. M., Jasilionis, D., Németh, L. et al. (2021). The short-term mortality fluctuations data series, monitoring mortality shocks across time and space. *Sci Data*, 8, Article 235. https://www.nature.com/articles/s41597-021-01019-1

2. Preston, S. H. & Vierboom, Y. C. (2021). Excess mortality in the United States in the 21^st^ century. *Proc Natl Acad Sci U S A*., 118, Article e2024850118. https://www.pnas.org/doi/10.1073/pnas.2024850118

3. Caporali, A., Garcia, J., Couppié, É., Poniakina, S., Barbieri, M., Bonnet, F. et al. (2022). The demography of COVID-19 deaths database, a gateway to well-documented international data. *Sci Data*, 9, Article 93. https://www.nature.com/articles/s41597-022-01191-y

4. NCHS. Provisional COVID-19 deaths by age and sex. https://data.cdc.gov/NCHS/Provisional-COVID-19-Death-Counts-by-Sex-Age-and-S/9bhg-hcku

5. United Nations. World Population Prospects 2019, online edition. <https://population.un.org/wpp/Download/Standard/Interpolated/>

6. Johns Hopkins University. Coronavirus Resource Center. https://coronavirus.jhu.edu
